# Assessment of Undergraduate Psychological Wellbeing Using Explainable Machine Learning: A Study from Sri Lanka

**DOI:** 10.64898/2026.11.10.26367798

**Authors:** Githmi Paranawithana, Anoma Perera, Shalika Kumburegama, Poornika Seelagama, Kanthi Hettigoda, Thushara Kamalrathne, Asanka Bulathwatta, Damayanthi Herath, Sayuri Perera, Ruchika Fernando

## Abstract

Academic, psychosocial, financial, environmental, and interpersonal factors are widely recognized as important contributors to the overall wellbeing of students. This study aimed to identify factors associated with the psychological wellbeing of university undergraduates and compare the performance of machine learning models for binary and multiclass classification of wellbeing status using self-reported survey data from undergraduates at the University of Peradeniya, Sri Lanka. The findings may facilitate the early identification of students with lower wellbeing and inform targeted interventions to foster a healthier and more supportive academic environment. Cross-sectional questionnaire data from 2,913 undergraduates were analyzed to classify psychological wellbeing using overall happiness scores. Scores were grouped into binary (1-5 and 6-9) and multiclass (1-4, 5-6, and 7-9) categories. Data preprocessing included missing value imputation, one-hot encoding, and feature scaling where appropriate. Support Vector Machine (SVM), Random Forest (RF), XGBoost (XGB), and Multilayer Perceptron (MLP) models were trained using repeated stratified 10-fold cross-validation and evaluated using macro F1-score, ROC-AUC, and classification reports. SHAP analysis was employed to interpret feature contributions and enhance model interpretability. XGB achieved the highest macro-F1 score (70.5%) for binary classification, with a test accuracy of 72.5%. The model correctly classified 63.6% and 80% of participants in the lower and higher psychological wellbeing groups, respectively. For multiclass classification, RF achieved the highest macro-F1 score (54.6%), with a test accuracy of 56.5%. The model correctly classified 48%, 57%, and 60% of participants in the lower, intermediate, and higher psychological wellbeing groups, respectively. SHAP analysis identified academic coping, health and wellbeing, financial circumstances, and interpersonal relationships as key predictors of psychological wellbeing across both models. These findings demonstrate the potential of machine learning to classify psychological wellbeing groups using self-reported data, supporting early identification of students at risk of poor wellbeing and informing targeted university-level mental health interventions.

**Author summary:** Psychological wellbeing among university students is influenced by various factors, including academic experiences, financial circumstances, health, social relationships, and available support. Identifying students with lower psychological wellbeing can help universities develop appropriate support programmes. This study explored the use of machine learning to classify psychological wellbeing among undergraduate students at the University of Peradeniya, Sri Lanka.

Data were collected through self-reported questionnaires completed by 2,913 undergraduates. Four machine learning models were developed and compared to classify students into two or three wellbeing categories. Explainable machine learning techniques were also applied to identify factors contributing most strongly to model predictions.

XGBoost performed best in classifying students into two wellbeing categories, while Random Forest performed best in distinguishing three wellbeing levels. The ability to cope with academic workload, satisfaction with personal health, social relationships, financial circumstances, and availability of non-financial support emerged as important predictors.

These findings highlight the potential of machine learning to identify patterns associated with psychological wellbeing and inform targeted university support programmes. However, such models should complement, rather than replace, professional assessment and established student support services.

## 1. Introduction

Mental wellbeing of students is a major public health and educational concern as it influences academic achievement, productivity, social functioning, retention, and long-term health outcomes. Student psychological wellbeing is shaped by complex interactions among academic, social, financial, environmental, and personal factors rather than by any single factor alone [1, 2].

Increased academic demands, changing social environments, financial pressures, reduced access to familiar support networks, and uncertainty about future careers may contribute to psychological distress and lower psychological wellbeing among undergraduates [3]. University students often report poorer wellbeing than their non-student peers of a similar age, potentially due to the unique demands associated with higher education [4–6]. The transition from secondary to tertiary education is a significant period of adjustment during which students may face academic, social, and financial challenges that can influence their mental health and psychological wellbeing [3,7].

Supporting student wellbeing is an important priority in Sri Lanka, where university students may face barriers to accessing psychological support, including limited service accessibility and social stigma associated with psychiatric treatment and mental health care. Despite growing awareness of mental health challenges, research investigating wellbeing among Sri Lankan undergraduates remains scanty. During the COVID-19 pandemic, a survey of 637 second-year students from three faculties at the University of Jaffna found that 31% screened positive for major depressive disorder, while nearly 70% reported varying levels of depressive symptoms, emphasizing the vulnerability of university populations in the country [8]. A study involving 100 students at the University of Ruhuna found that academic, financial, social, environmental, institutional, and accommodation-related factors were associated with psychiatric symptoms among undergraduates [9].

Traditional statistical approaches in student wellbeing research, including regression-based models and descriptive analyses are frequently used [10]. However, these conventional analytical approaches may inadequately capture complex nonlinear relationships, interactions among predictors, overlapping classes, and multidimensional dependencies in behavioral and psychosocial datasets. Machine learning methods offer alternative analytical frameworks capable of modeling these complex relationships while improving predictive performance and supporting early identification of vulnerable populations. Recent studies have demonstrated the usefulness of machine learning approaches for predicting mental wellbeing using behavioral, academic, and health-related data, with algorithms such as Random Forest, Support Vector Machines, Artificial Neural Networks, and gradient boosting methods demonstrating promising predictive capability [11, 12].

Therefore, this study aimed to identify factors associated with the psychological wellbeing of undergraduates at the University of Peradeniya, Sri Lanka and compare the performance of machine learning models in classifying wellbeing status using binary and multiclass approaches based on self-reported survey data. By integrating predictive modelling with explainable machine learning techniques, this study sought to elucidate the relative contributions of multidimensional factors associated with psychological wellbeing and explore the potential of these models to support the early identification of students at risk of poor wellbeing and inform targeted university-level interventions.

## 2. Materials and methods

### 2.1 Ethics statement

Ethical approval for this study was obtained from the Institutional Ethics Review Committee of the Faculty of Arts, University of Peradeniya, Sri Lanka prior to data collection (Approval No: Arts/Ethics/2024/01/20.1). Participation in the study was voluntary, and written informed consent was obtained from all participants before enrolment. Confidentiality and anonymity of the collected information were strictly maintained throughout the research process.

### 2.2 Study design

A questionnaire-based cross-sectional analytical study was conducted among undergraduates of the University of Peradeniya from 06.03.2024 to 31.07.2025 to evaluate factors associated with their psychological wellbeing and self-reported happiness. The survey responses were used to apply machine learning approaches for classifying participants into wellbeing categories. Each participant completed the survey only once.

### 2.3 Study population and eligibility

This study included undergraduate students enrolled in degree programmes across seven faculties of the University: Agriculture, Allied Health Sciences, Arts, Engineering, Management, Science, and Veterinary Medicine and Animal Science. Participants were selected using stratified random sampling to ensure representation proportional to student enrolment across faculties and academic years. However, in the Faculty of Medicine, only first- and fifth-year undergraduates were accessible for participation in the survey. Written informed consent was obtained from the participants. Students who were receiving or had previously received long-term treatment, including medication, for chronic physical or mental health conditions were excluded from the survey. After eligibility assessment and data processing, responses from 2,913 participants were included in the present analysis.

### 2.4 Data Collection Instrument

Data were collected using a questionnaire developed and validated by the research team to assess demographic, financial, interpersonal and psychosocial, environmental, and academic characteristics relevant to student wellbeing and self-reported happiness. All collected data were de-identified prior to analysis to ensure participant confidentiality and anonymity.

### 2.5 Dataset and predictor variables

Table 1 summarizes the predictor variables extracted from the data collection instrument for the analysis. Detailed descriptions of the predictors, including their operational definitions, coding schemes, and measurement scales, are provided in <u>Table A of S1 Text.</u>

**Table 1.** Predictor Variables Extracted from the Data Collection Instrument.

| Variable Category | Predictor Variable |
|---|---|
| Demographic variables | Faculty, academic year, sex, religion |
| Financial variables | Income, money_happiness |
| Interpersonal and psychosocial variables | nonFinancialHelp, decisionFreedom, goalsAchieve, relationships, acceptance, trusted, getHelp, suspected, disliked, happyRelationships, loveRelationships, kindRelationships, fearfulRelationships, discrimRelationships, healthSatisfaction |
| Environmental variables | facOverallEnv, facNatuEnv, livingArrangement, hostelOverallEnv, hostelEnv, homeOverallEnv |
| Academic variables | academicTolerance, GPA |

### 2.6 Target Variable/Outcome

The target variable in this study was the *Happiness Group*, representing students’ self-reported overall happiness assessments. This was measured using a nine-point Likert scale, where a score of 1 represented *extreme unhappiness,* and a score of 9 represented *extreme happiness*. For binary classification, overall happiness scores were grouped into two groups: Group 1 (lower wellbeing; scores 1-5) and Group 2 (higher wellbeing; scores 6-9). For multiclass classification, scores were categorized into three groups: Group 1 (lower wellbeing; scores 1-4), Group 2 (intermediate wellbeing; scores 5-6), and Group 3 (higher wellbeing; scores 7-9). This approach enabled a more detailed assessment of wellbeing categories using students’ self-reported happiness scores.

### 2.7 Modeling software

Data preprocessing, model development, hyperparameter tuning, model evaluation, and statistical analyses were performed using Python within the Google Colaboratory environment. Data manipulation and preprocessing procedures were conducted using *pandas* and *NumPy*, while all machine learning models, including Extreme Gradient Boosting (XGBoost), Random Forest (RF), Support Vector Machine (SVM), Naive Bayes (NB), and Multilayer Perceptron (MLP) were implemented using Python and their respective machine learning libraries. Model interpretability analyses were conducted using the SHAP (SHapley Additive exPlanations) library. Additional visualization and data exploration procedures were performed using Python-based analytical tools within the same computational environment to ensure reproducibility and scalability of analyses.

### 2.8 Data preprocessing

Initially, the dataset was examined for missing values and inconsistencies. Missing data were handled using imputation techniques to preserve the overall sample size and reduce information loss. The target variable contained no missing values. Missing values in the encoded predictor variables were handled using variable-wise median imputation. Several preprocessing procedures were applied to improve data quality and compatibility with machine learning algorithms. Categorical variables, including gender identity and residential status, were transformed using one-hot encoding to convert them into machine readable binary representations. Feature scaling was applied using standardization techniques for models sensitive to feature magnitudes, particularly Multilayer Perceptrons (MLPs), linear SVMs, and non-linear SVMs [12]. Following one-hot encoding, a correlation matrix was generated to assess multicollinearity among predictors. Feature pairs with absolute correlation coefficients ≥0.70 were examined for potential redundancy, and features below this threshold were retained for subsequent feature selection. Feature selection was conducted using the Boruta algorithm to identify all relevant predictors associated with the classification of psychological wellbeing status of the participants. The robustness of the selected features was assessed through a stability analysis by repeating the Boruta feature selection process across 30 randomized subsamples of the dataset. A feature was retained only if it was identified as “important” in at least 70% of the iterations. Furthermore, the stability of the selected predictors was validated using model-based feature importance rankings derived from the trained machine learning classifiers (<u>Fig. A & B in</u> <u>S1 Text</u>). Independent variables measured on the 9-point Likert scale were categorized into three ordinal groups (1-4, 5-6, and 7-9). Goodman and Kruskal’s gamma coefficients were then calculated to assess the strength and direction of ordinal associations between each selected independent variable and the dependent variable (multiclass wellbeing group).

### 2.9 Machine learning algorithms and performance evaluation

The dataset was divided into training and testing subsets using a stratified split, where 70% of the data were allocated for model training and 30% reserved for testing. The target classes exhibited moderate imbalance in both the binary and multiclass classification tasks. Therefore, stratification was performed to ensure proportional distribution of the two and three wellbeing classes. To further evaluate model robustness and reduce sampling variability, repeated 10-fold stratified cross validation was employed during model development and hyperparameter tuning. Multiple machine learning algorithms, including RF, NB, XGBoost, linear and non-linear SVM, and MLPs were trained and compared within the preprocessing and modeling framework. Together, these preprocessing procedures ensured that the dataset was appropriately structured, normalized, and validated for robust binary and multiclass wellbeing classification analysis. The machine learning workflow of the study is demonstrated in Fig 1.

**Fig 1.**
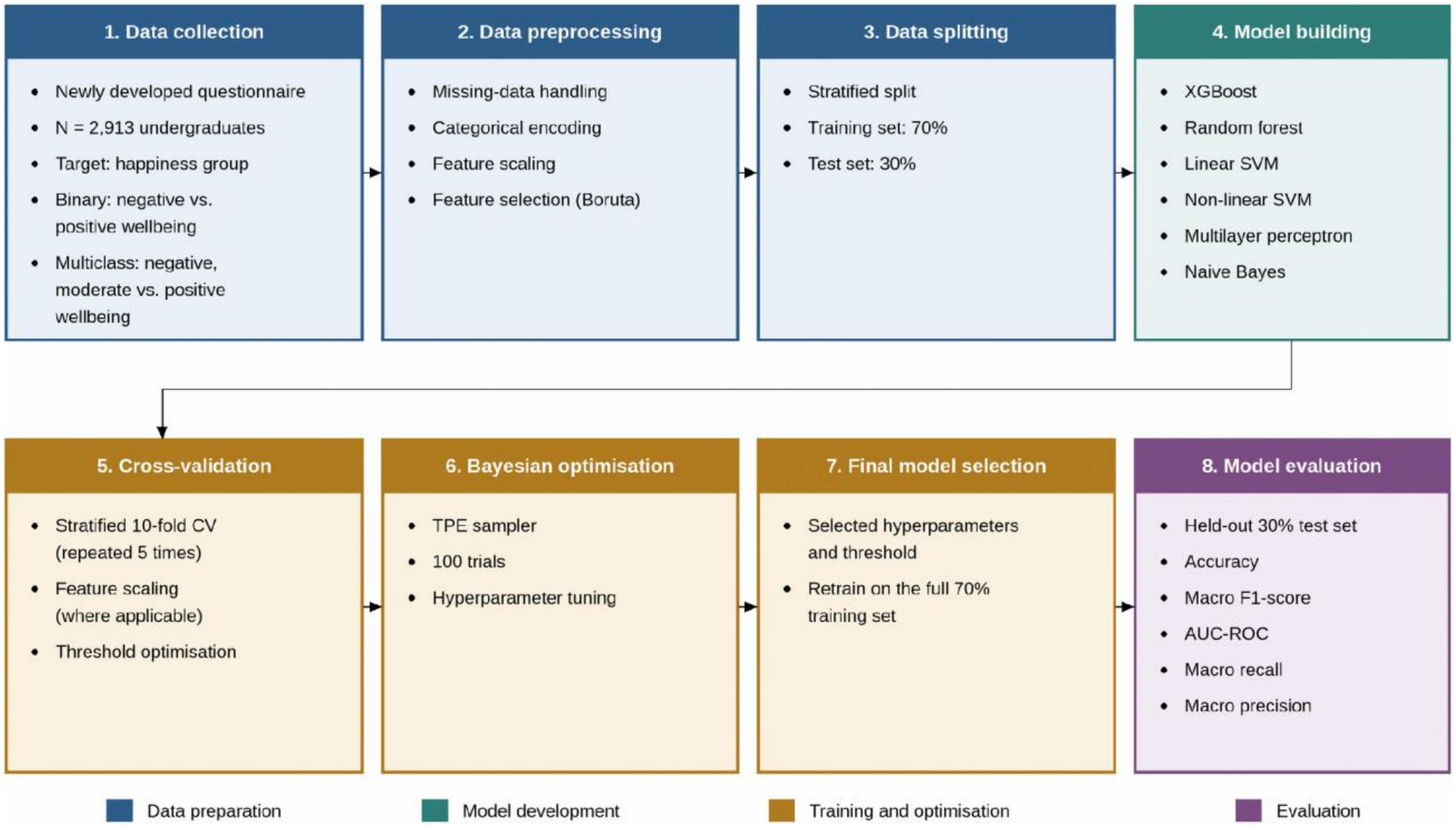
Machine Learning Workflow for Classification of Psychological Wellbeing Status Among Undergraduates of the University of Peradeniya.

Model performance was evaluated using multiple metrics, including cross validation accuracy, test accuracy, macro F1-score, classification reports, and normalized confusion matrices. Due to the presence of moderate class imbalance across wellbeing groups, macro averaged metrics were prioritized to ensure balanced performance assessment across categories. This multi-model framework enabled comprehensive comparison across fundamentally different learning approaches while supporting identification of the most reliable and interpretable predictive models for student wellbeing classification.

### 2.10 Hyperparameter tuning and model building

Hyperparameter optimization was performed separately for the binary and multiclass classification tasks to accommodate their distinct prediction objectives and class structures. For each machine learning algorithm, Bayesian optimization using the Tree-structured Parzen Estimator (TPE) was employed to identify optimal hyperparameter configurations within predefined search spaces. The hyperparameters optimized for the binary and multiclass classification tasks are presented in [Table B](https://docs.google.com/document/d/1oB3DtT84XhLvibKNwuXBlytn0ZQXuXi-P33ZEdqL9ag/edit?tab=t.0#bookmark=id.f8xao9x2c9qr) and [Table C of the S1 Text](https://docs.google.com/document/d/1oB3DtT84XhLvibKNwuXBlytn0ZQXuXi-P33ZEdqL9ag/edit?tab=t.0#bookmark=id.a6bclebntzqo), respectively.

### 2.11 Model interpretability

SHAP analysis was used separately for binary and multiclass wellbeing classifications to interpret model predictions and identify the most influential demographic, financial, interpersonal and psychosocial, environmental, and academic determinants. For each classification task, SHAP analyses were performed on the best-performing model to identify the relative importance and direction of feature contributions. SHAP beeswarm and bar plots were generated to visualize the influence of feature values on model predictions towards the respective wellbeing categories and summarize global feature importance based on the mean absolute SHAP values, respectively.

## 3. Results

### 3.1 Sociodemographic characteristics

<u>Table D in S1 Text</u> provides background characteristics of the binary classification of overall wellbeing of the participants. 979 (33.61%) and 1,934 (66.39%) participants were classified into the lower and higher wellbeing categories, respectively, indicating a predominance of higher wellbeing in the study population. In the multiclass classification framework, 510 (17.51%), 1,148 (39.41%), and 1,255 (43.08%) participants were classified into the lower, intermediate, and higher wellbeing categories, respectively <u>(Table E in S1 Text</u>). Overall, higher happiness scores (6-9) were more frequently reported than lower scores (1-4) in all gender categories (Table 2). In the multiclass classification of student overall wellbeing, Goodman and Kruskal’s Gamma analysis revealed statistically significant positive ordinal associations between all investigated independent variables measured on a 9-point Likert scale and undergraduate wellbeing categories (all p < 0.001) (Table 3).

**Table 2:** Distribution of Overall Happiness scores by gender among undergraduate students.

| **Gender** | **Overall Happiness Score** |  |  |  |  |  |  |  |  |
|---|---|---|---|---|---|---|---|---|---|
|  | **1** | **2** | **3** | **4** | **5** | **6** | **7** | **8** | **9** |
| **Female (n = 2107)** | 46 (2.19) | 61 (2.90) | 65 (3.09) | 192 (9.12) | 324 (15.39) | 475 (22.57) | 573 (27.22) | 225 (10.69) | 144 (6.84) |
| **Male (n = 726)** | 22 (2.76) | 20 (2.51) | 27 (3.39) | 76 (9.55) | 143 (17.96) | 203 (25.50) | 196 (24.62) | 67 (8.42) | 42 (5.28) |
| **Not disclosed (n = 10)** | 0 (0.00) | 0 (0.00) | 1 (10.00) | 0 (0.00) | 2 (20.00) | 1 (10.00) | 4 (40.00) | 1 (10.00) | 1 (10.00) |

**Note:** Percentages within gender categories are presented in parentheses.

**Table 3:** Distribution of 9-point Likert scale-based independent variables across the three wellbeing groups among undergraduate students.

| Feature | Likert Category | Group 1: Lower Wellbeing Frequency (%) | Group 2: Intermediate Wellbeing Frequency (%) | Group 3: Higher Wellbeing Frequency (%) | Goodman and Kruskal's Gamma (γ) | p-value |
|---|---|---|---|---|---|---|
| **Money_happiness** | 1-4 | 337 (24.9) | 587 (43.4) | 430 (31.8) | 0.377 | p < 0.001 |
|  | 5-6 | 120 (12.8) | 392 (41.9) | 424 (45.3) |  |  |
|  | 7-9 | 53 (8.5) | 169 (27.1) | 401 (64.4) |  |  |
| **nonFinancialHelp** | 1-4 | 219 (32.5) | 267 (39.6) | 188 (27.9) | 0.416 | p < 0.001 |
|  | 5-6 | 156 (18.1) | 432 (50.2) | 272 (31.6) |  |  |
|  | 7-9 | 135 (9.8) | 449 (32.6) | 795 (57.7) |  |  |
| **decisionFreedom** | 1-4 | 96 (32.9) | 128 (43.8) | 68 (23.3) | 0.326 | p < 0.001 |
|  | 5-6 | 139 (20.1) | 324 (47.0) | 227 (32.9) |  |  |
|  | 7-9 | 275 (14.2) | 696 (36.0) | 960 (49.7) |  |  |
| **goalsAchieve** | 1-4 | 92 (52.6) | 58 (33.1) | 25 (14.3) | 0.473 | p < 0.001 |
|  | 5-6 | 173 (22.9) | 378 (50.1) | 203 (26.9) |  |  |
|  | 7-9 | 245 (12.3) | 712 (35.9) | 1,027 (51.8) |  |  |
| **relationships** | 1-4 | 119 (51.5) | 78 (33.8) | 34 (14.7) | 0.505 | p < 0.001 |
|  | 5-6 | 200 (22.3) | 447 (49.8) | 250 (27.9) |  |  |
|  | 7-9 | 191 (10.7) | 623 (34.9) | 971 (54.4) |  |  |
| **healthSatisfaction** | 1-4 | 215 (39.0) | 211 (38.3) | 125 (22.7) | 0.460 | p < 0.001 |
|  | 5-6 | 189 (17.3) | 517 (47.3) | 388 (35.5) |  |  |
|  | 7-9 | 106 (8.4) | 420 (33.1) | 742 (58.5) |  |  |
| **facOverallEnv** | 1-4 | 116 (30.0) | 160 (41.3) | 111 (28.7) | 0.278 | p < 0.001 |
|  | 5-6 | 186 (17.9) | 480 (46.1) | 376 (36.1) |  |  |
|  | 7-9 | 208 (14.0) | 508 (34.2) | 768 (51.8) |  |  |
| **facNatuEnv** | 1-4 | 45 (33.3) | 48 (35.6) | 42 (31.1) | 0.218 | p < 0.001 |
|  | 5-6 | 115 (19.2) | 282 (47.1) | 202 (33.7) |  |  |
|  | 7-9 | 350 (16.1) | 818 (37.5) | 1,011 (46.4) |  |  |
| **hostelOverallEnv** | 1-4 | 235 (25.4) | 387 (41.8) | 304 (32.8) | 0.283 | p < 0.001 |
|  | 5-6 | 160 (15.9) | 445 (44.2) | 401 (39.9) |  |  |
|  | 7-9 | 115 (11.7) | 316 (32.2) | 550 (56.1) |  |  |
| **hostelEnv** | 1-4 | 194 (26.7) | 290 (39.9) | 242 (33.3) | 0.262 | p < 0.001 |
|  | 5-6 | 140 (15.6) | 432 (48.1) | 327 (36.4) |  |  |
|  | 7-9 | 176 (13.7) | 426 (33.1) | 686 (53.3) |  |  |
| **homeOverallEnv** | 1-4 | 42 (37.8) | 43 (38.7) | 26 (23.4) | 0.321 | p < 0.001 |
|  | 5–6 | 87 (22.9) | 183 (48.2) | 110 (28.9) |  |  |
|  | 7-9 | 381 (15.7) | 922 (38.1) | 1,119 (46.2) |  |  |
| **academicTolerance** | 1-4 | 274 (37.2) | 268 (36.4) | 194 (26.4) | 0.456 | p < 0.001 |
|  | 5-6 | 165 (13.1) | 610 (48.4) | 485 (38.5) |  |  |
|  | 7-9 | 71 (7.7) | 270 (29.4) | 576 (62.8) |  |  |

### 3.2 Hyperparameter tuning and training performance

The hyperparameter tuning process identified the optimal configurations for each machine learning model in the binary and multiclass classification tasks. The hyperparameter search spaces explored and the corresponding optimal values for each model are presented in [Tables B](https://docs.google.com/document/d/1oB3DtT84XhLvibKNwuXBlytn0ZQXuXi-P33ZEdqL9ag/edit?tab=t.0#bookmark=id.f8xao9x2c9qr) and [C](https://docs.google.com/document/d/1oB3DtT84XhLvibKNwuXBlytn0ZQXuXi-P33ZEdqL9ag/edit?tab=t.0#bookmark=id.a6bclebntzqo) in S1 Text for the binary and multiclass classification frameworks, respectively. For the binary classification task, the performance of each machine learning model is indicated in Table 4. The optimized XGBoost (XGB) model achieved the strongest overall performance based on all considered evaluation metrics.

**Table 4.** Performance evaluation of machine learning models for binary classification using cross validation on the training dataset.

| **ML Model** | **Accuracy (95% CI)** | **Macro-Recall** | **Macro-Precision** | **AUC-ROC** | **Macro-F1** |
|---|---|---|---|---|---|
| XGBoost | 0.7504 (0.7322, 0.7690) | 0.7276 | 0.7215 | 0.8005 | 0.7242 |
| Random Forest | 0.7308 (0.7014, 0.7602) | 0.7244 | 0.7074 | 0.7938 | 0.7117 |
| Linear SVM | 0.7018 (0.6817, 0.7214) | 0.7095 | 0.6981 | 0.7730 | 0.7021 |
| Non-linear SVM | 0.7258 (0.7067, 0.7450) | 0.7240 | 0.7050 | 0.7801 | 0.7086 |
| Naive Bayes | 0.7249 (0.7057, 0.7445) | 0.7135 | 0.6992 | 0.7802 | 0.7034 |
| MLP | 0.7322 (0.7131, 0.7513) | 0.7133 | 0.7036 | 0.7783 | 0.7073 |

For the multiclass classification task, the optimized RF model demonstrated the strongest overall classification performance, achieving an accuracy of 60.0% (95% CI: 57.9%, 62.1%), macro-recall of 59.0%, macro-precision of 58.9%, macro-F1-score of 58.8%, and macro AUC-ROC (Area Under the Receiver Operating Characteristic Curve) of 76.7%. RF and XGB achieved the highest accuracy and macro-F1 scores, while NB demonstrated intermediate performance. Linear SVM, non-linear SVM, and MLP yielded comparatively lower macro-F1 scores (Table 5). These findings indicate that the ensemble-based models, particularly RF and XGB, provided the most favorable overall performance for distinguishing among the three wellbeing outcome categories.

**Table 5.** Performance evaluation of machine learning models for multiclass classification using cross validation on the training dataset.

| **ML Model** | **Accuracy (95% CI)** | **Macro-Recall** | **Macro-Precision** | **Macro AUC-ROC** | **Macro-F1** |
|---|---|---|---|---|---|
| XGBoost | 0.5998 (0.5787, 0.6209) | 0.5833 | 0.5842 | 0.7686 | 0.5837 |
| Random Forest | 0.5998 (0.5787, 0.6214) | 0.5904 | 0.5891 | 0.7665 | 0.5882 |
| Linear SVM | 0.5677 (0.5581, 0.5775) | 0.5526 | 0.5573 | 0.7319 | 0.5524 |
| Non-linear SVM | 0.5623 (0.5526, 0.5716) | 0.5553 | 0.5465 | 0.7344 | 0.5470 |
| Naive Bayes | 0.5717 (0.5621, 0.5813) | 0.5599 | 0.5700 | 0.7497 | 0.5596 |
| MLP | 0.5660 (0.5440, 0.5880) | 0.5601 | 0.5389 | 0.7294 | 0.5292 |

### 3.3 Performance on the test set

Fig. 2 presents the performance metrics of the six machine learning models on the independent test set used for the binary classification of psychological wellbeing outcomes. The models were primarily evaluated based on macro F1-score and ROC-AUC.

**Fig 2.**
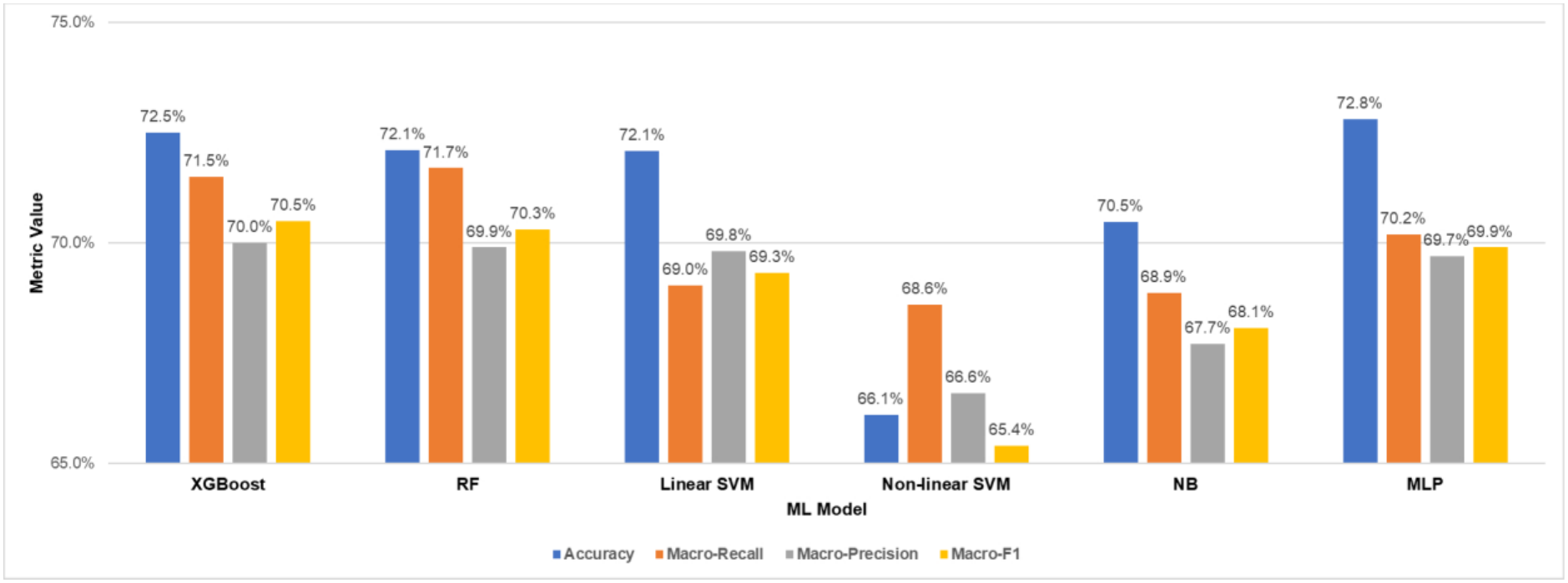
Performance of machine learning models on the test set for binary classification of self-reported happiness among undergraduates of the University of Peradeniya.

Among the evaluated models, XGB demonstrated the strongest overall binary classification performance, achieving the highest test set macro F1-score (71.6%) and macro-recall (71.8%), together with a test accuracy of 74.5% (95% CI: 71.5, 77.4%) and a ROC-AUC of 79.4%. Its macro-precision was 71.5%, indicating relatively balanced predictive performance across the lower and higher wellbeing categories despite the moderate class imbalance. These findings suggest that XGB provided the most favorable balance between overall classification performance and sensitivity across the two classes. The linear SVM model showed the lowest macro F1-score. Although MLP yielded the highest accuracy, its lower sensitivity illustrates why accuracy alone may be insufficient for model comparison in the presence of class imbalance. XGB further offered a more favorable balance between macro-averaged performance and class-specific sensitivity, whereas RF demonstrated particularly balanced sensitivity between the two wellbeing categories.

Fig. 3 presents the performance metrics of the six machine learning models on the independent test set used for the multiclass classification of psychological wellbeing outcomes. The models were primarily evaluated based on macro F1-score and ROC-AUC.

**Fig 3.**
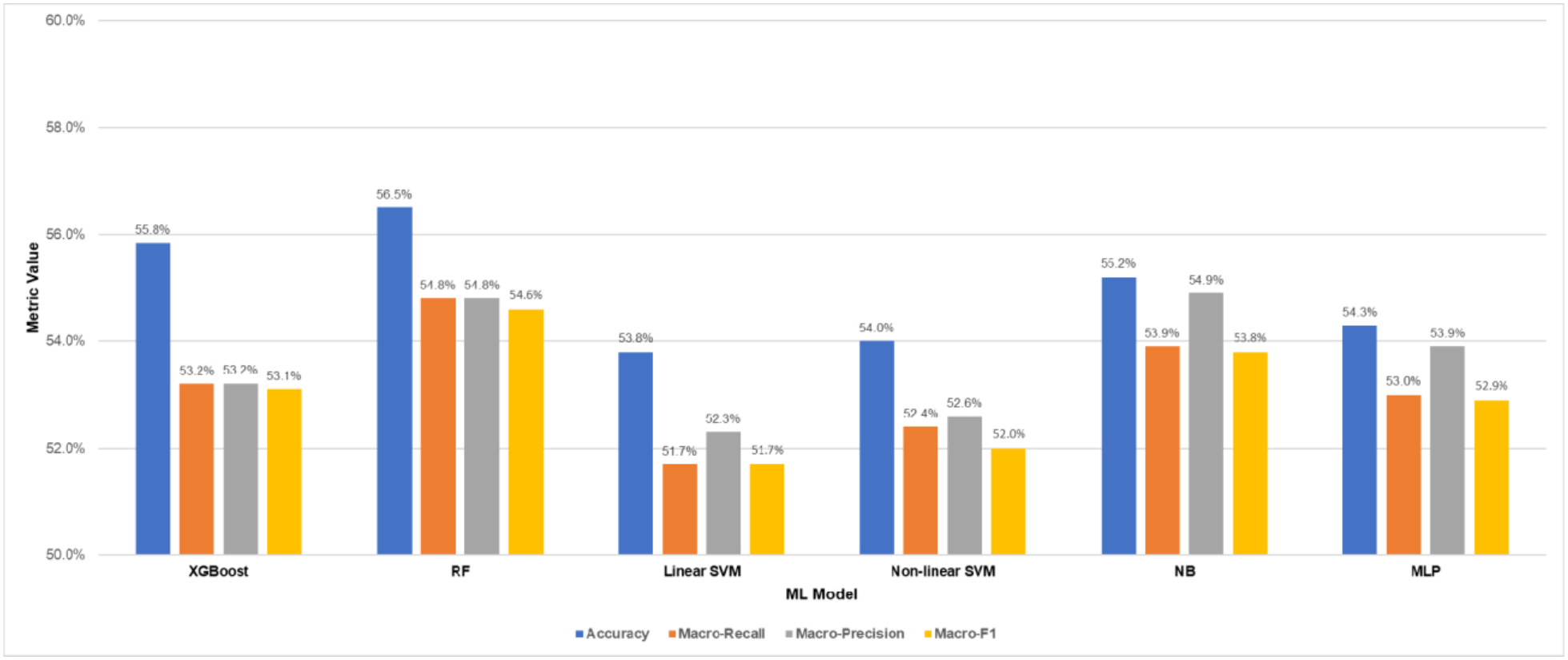
Performance of machine learning models on the test set for multiclass classification of self-reported overall happiness among undergraduates of the University of Peradeniya.

Overall, RF achieved the strongest multiclass classification performance in terms of accuracy and macro-averaged classification metrics, while XGB achieved the highest AUC-ROC (73.9%). NB also demonstrated competitive performance, whereas the SVM-based models and MLP achieved comparatively lower macro-F1 scores. Across the six models, test accuracy ranged from 53.8% to 56.5%, macro-recall from 51.7% to 54.8%, macro-precision from 52.3% to 54.9%, macro-F1 from 51.7% to 54.6%, and AUC-ROC from 70.8% to 73.9%. These findings indicate a moderate ability to distinguish among lower, intermediate, and higher wellbeing categories. Compared with binary classification, the multiclass classification task requires models to differentiate among a greater number of outcome categories, including the intermediate wellbeing group.

### 3.4 Model interpretability

Fig 4(a) and Fig 4(b) show SHAP bar and beeswarm plots for the binary XGB model. In binary classification, academic tolerance emerged as the most influential predictor, with a mean absolute SHAP value of 0.5263, followed by health satisfaction (0.4003), satisfaction regarding the amount of money they have for their needs (0.319), their relationships with others (0.299), overall environment of their hostels or boarding places (0.227), achievement of their goals (0.215), non-financial help received by others (0.197), and decision making freedom (0.094). The beeswarm distribution for academic tolerance showed that instances of higher levels of happiness were concentrated predominantly on the negative SHAP side, whereas lower values extended towards positive SHAP values. This pattern suggests that higher levels of academic tolerance and health satisfaction generally shifted model predictions towards higher wellbeing, while lower values tended to shift predictions towards lower wellbeing. Further, this pattern was observed across all features considered in the binary classification. The breadth of the distribution in the beeswarm plot highlights variability in the contribution of each variable across participants. The spread of points across both sides of zero in respective variables indicates variation in the magnitude and direction of individual contributions.

**Fig 4.**
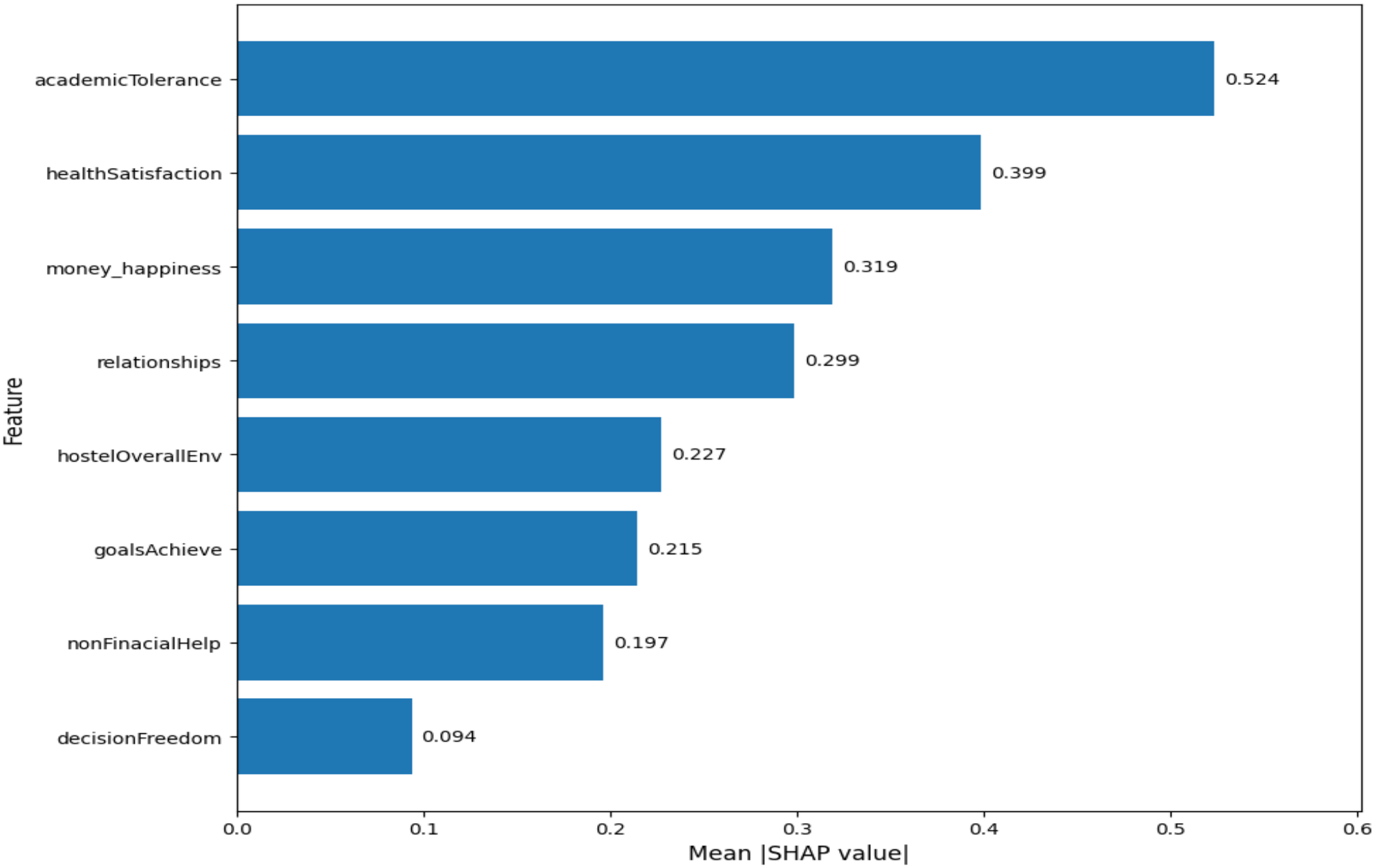
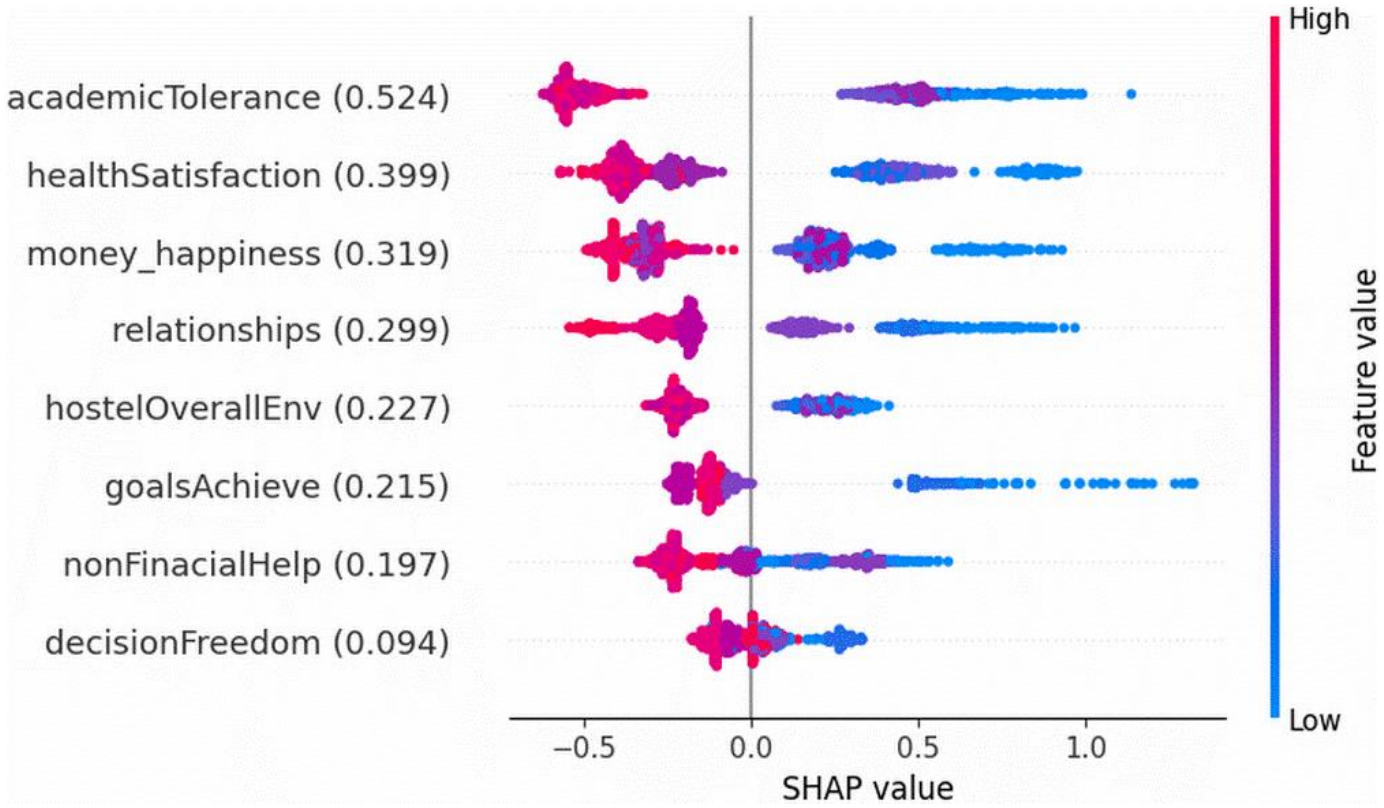
(a). SHAP Bar Plot of the XGB model for the binary classification of the Happiness Group of the undergraduates of the University of Peradeniya. (b). SHAP Beeswarm Plot of the XGB model for the binary classification of the Happiness Group of the undergraduates of the University of Peradeniya.

Overall, the SHAP analysis identified academicTolerance, healthSatisfaction, money_happiness, and relationships as the main contributors to the XGB model’s binary classification of undergraduate wellbeing. However, the distributions of SHAP values revealed variation across participants, indicating that predictions reflected the combined contributions of multiple features rather than the influence of any single predictor alone. These findings describe feature contributions within the fitted model and should not be interpreted as evidence of causal relationships.

For the multiclass Random Forest model, a SHAP stacked bar plot (Fig 5) and class-specific beeswarm plots (Fig 6 a-c) were generated to interpret predictions across the three wellbeing categories (lower, intermediate, and higher). These plots enabled the identification of predictors that were influential across the three wellbeing categories, as well as differences in the direction and magnitude of their contributions. The stacked bar plot summarized the summed mean absolute SHAP contributions of the seven selected features for each class, with the total contribution of the three classes used to compare their overall importance. Together, the two plots provided complementary insights into the global importance of the selected features and their class-dependent contributions to the model’s predictions.

**Fig 5.**
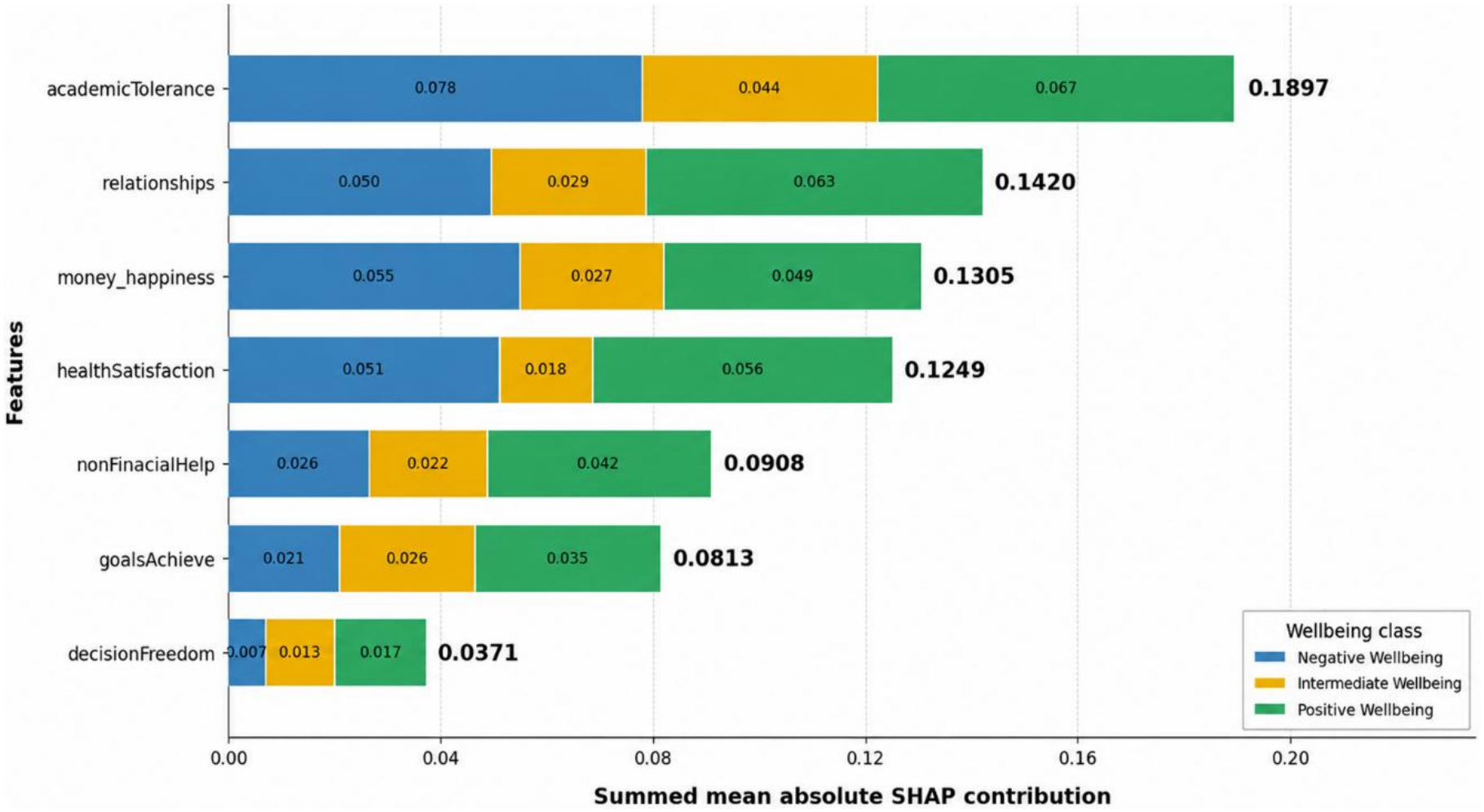
SHAP Stacked Bar Plot of the Random Forest model for the multiclass classification of the Happiness Group of the undergraduates of the University of Peradeniya.

**Fig 6.**
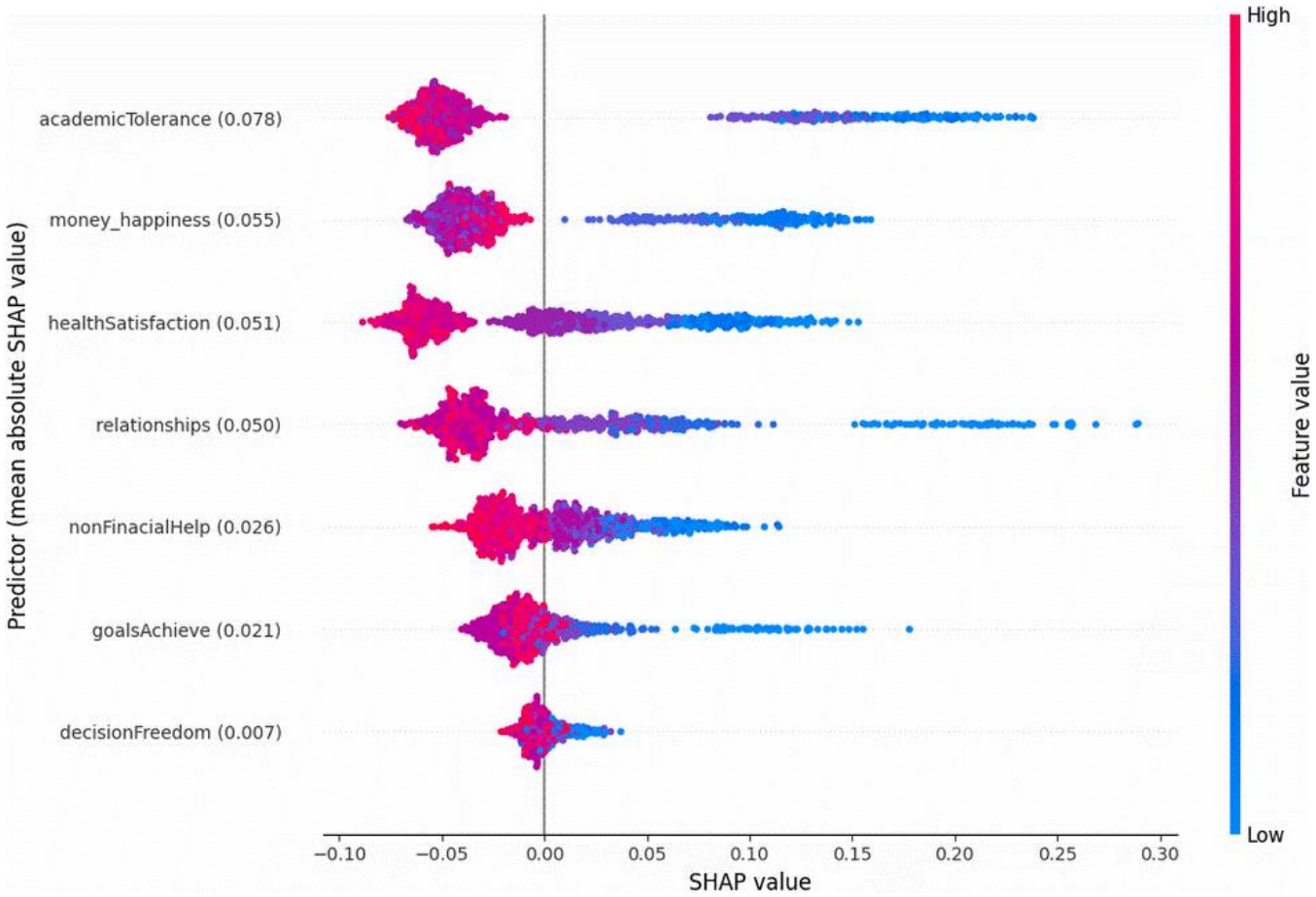
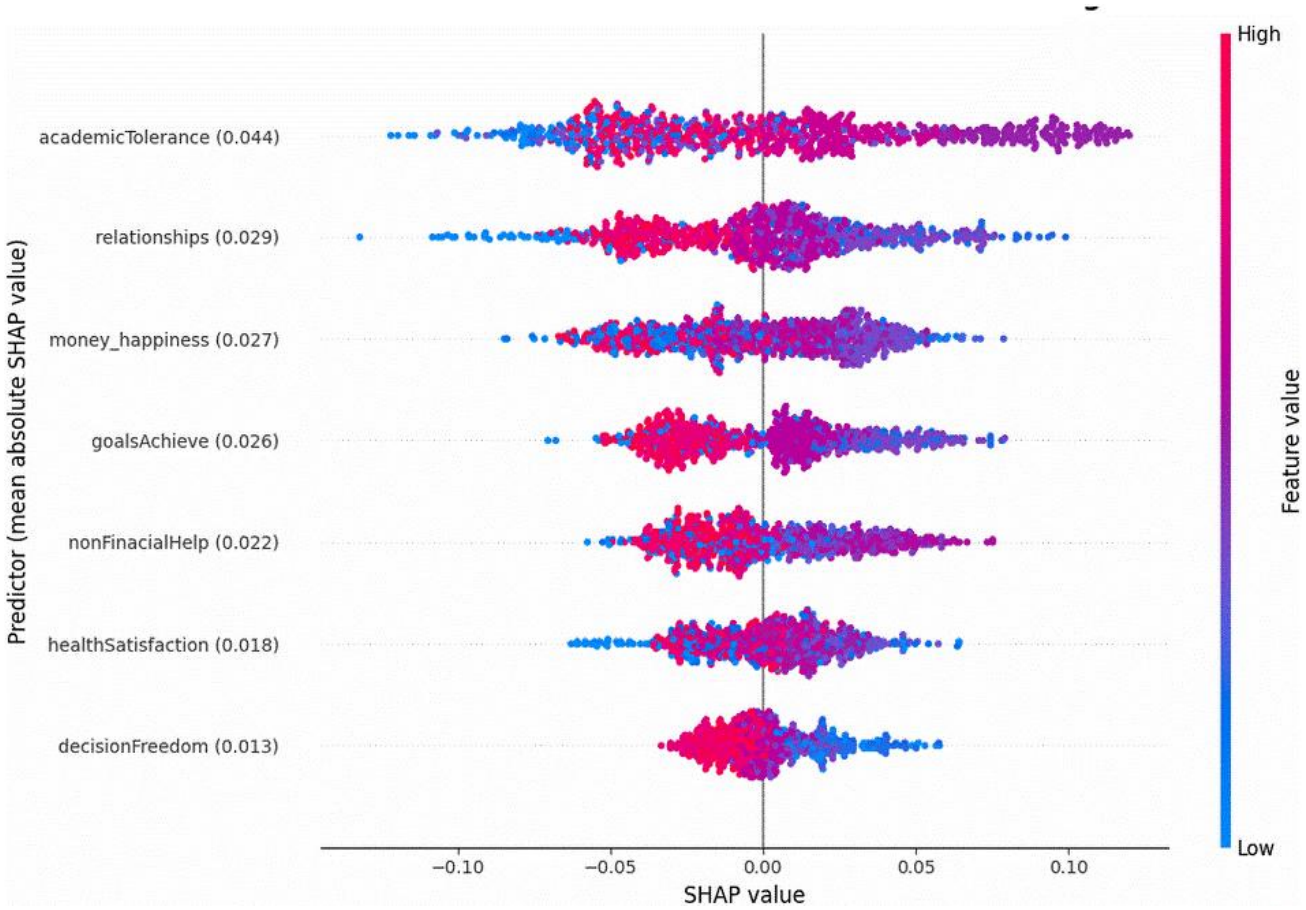
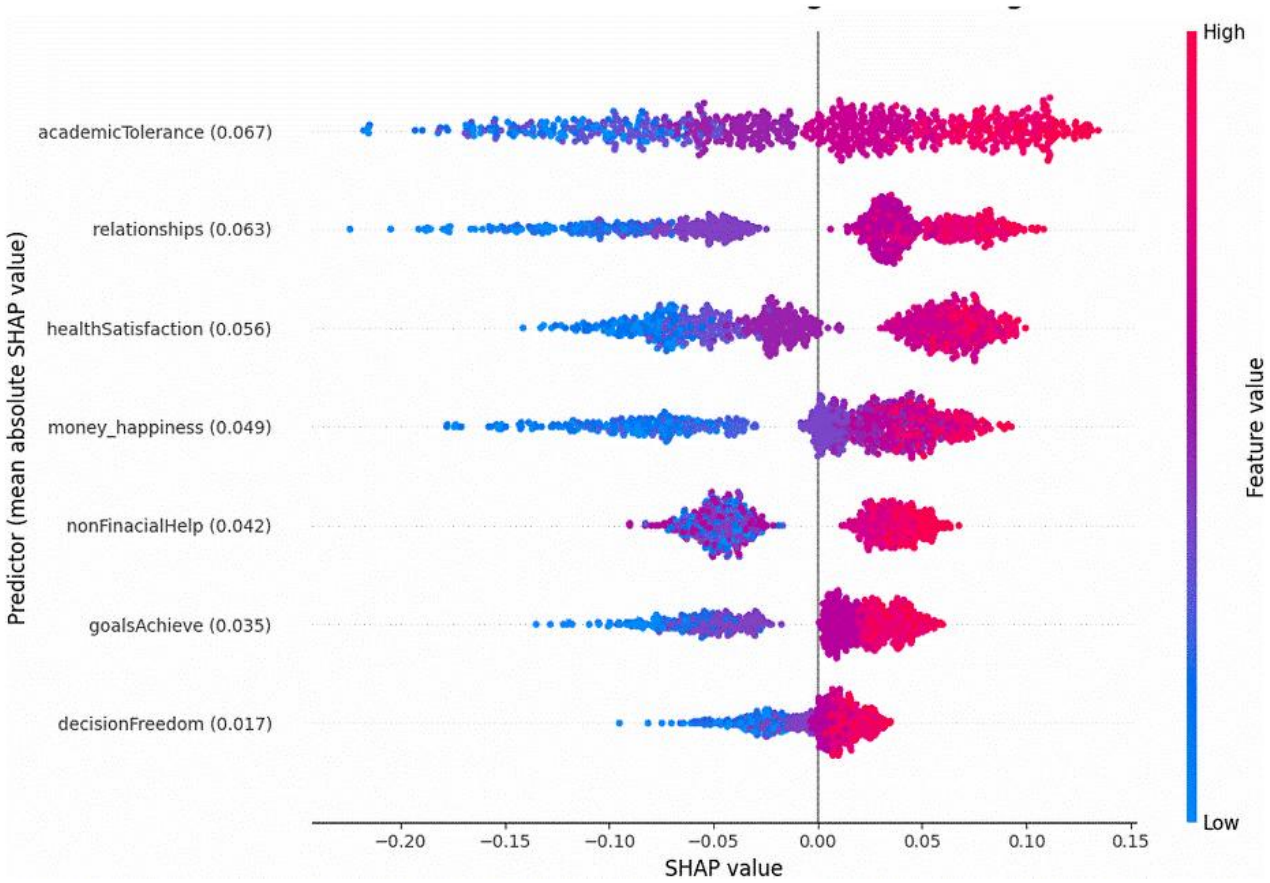
(a). SHAP Beeswarm Plot of the Random Forest Model for the Lower Wellbeing Category in the Multiclass Classification of Undergraduate Happiness Groups at the University of Peradeniya. (b). SHAP Beeswarm Plot of the Random Forest Model for the Intermediate Wellbeing Category in the Multiclass Classification of Undergraduate Happiness Groups at the University of Peradeniya. (c). SHAP Beeswarm Plot of the Random Forest Model for the Higher Wellbeing Category in the Multiclass Classification of Undergraduate Happiness Groups at the University of Peradeniya.

Consistent with the binary classification results, academic tolerance emerged as the most influential feature in the multiclass classification framework, exhibiting the highest mean absolute SHAP value (0.1897), with class-specific contributions of 0.078, 0.044, and 0.067 for lower, intermediate, and higher wellbeing, respectively, demonstrating its consistent importance in differentiating the three wellbeing categories. The students’ satisfaction regarding their relationships with others (0.1420), availability of money for their financial needs (0.1305), and their health and wellbeing (0.1249) were the next most influential features. Relationships contributed most strongly to higher wellbeing (0.063), whereas the students’ satisfaction regarding the availability of money for their financial needs showed its largest contribution to lower wellbeing (0.055). Health satisfaction exhibited relatively high contributions to higher (0.056) and lower wellbeing (0.051), but a lower contribution to intermediate wellbeing (0.018).

Non-financial help (0.0908) and goal achievement (0.0813) demonstrated intermediate levels of overall importance, whereas decision making freedom had the smallest total contribution (0.0371). Nevertheless, the contribution of each feature varied across the wellbeing categories, highlighting the importance of class-specific rather than exclusively aggregate interpretation. Overall, the results indicate that academic tolerance, interpersonal relationships, financial satisfaction, and health satisfaction were the principal features contributing to the Random Forest model’s classification decisions.

The class-specific beeswarm plots provided further insight into how each feature influenced the model’s predictions. For lower wellbeing, lower levels of academic tolerance, happiness with the availability of money to meet financial needs, health and wellbeing, and satisfaction with relationships generally contributed to a higher likelihood of the model predicting lower wellbeing. In contrast, higher values of these features generally contributed to predictions away from the lower wellbeing class. For higher wellbeing, higher levels of academic tolerance, relationship satisfaction, health satisfaction, happiness with financial resources, availability of non-financial help, and achievement of goals generally contributed to predictions of higher wellbeing, whereas lower values tended to contribute away from higher wellbeing. These findings indicate that the same features can influence the classification of different wellbeing categories in different ways, although the strength and direction of their contributions varied across individuals.

In contrast, the intermediate wellbeing class exhibited more heterogeneous SHAP distributions, with high and low feature values occurring on both sides of zero for several predictors. This pattern suggests that the model’s classification of intermediate wellbeing depends on a more variable combination of feature values than can be captured by a single directional association. Overall, the SHAP analysis identified academic tolerance as the dominant predictor across classes, while highlighting the complementary and class-dependent contributions of relationships, money_happiness, and healthSatisfaction. These findings describe the predictive behavior of the fitted RF model and should not be interpreted as evidence of causal relationships between the features and wellbeing categories.

## 4. Discussion

This study aimed to identify factors associated with the psychological wellbeing of undergraduates at the University of Peradeniya, Sri Lanka, and compare the performance of machine learning models in classifying wellbeing status using binary and multiclass approaches based on self-reported survey data. By integrating predictive modelling with explainable machine learning techniques, this study sought to elucidate the relative contributions of a number of selected multidimensional factors associated with psychological wellbeing and explore the potential of these models to support the early identification of students at risk of poor wellbeing and inform targeted university-level interventions.

The study identified academic, interpersonal and psychosocial, and financial factors as the most affecting factors for the wellbeing levels derived from the self-reported overall happiness of the undergraduates. Out of the two academic factors assessed, the students’ ability to cope with their academic workload was found to be the most influential factor contributing to the overall wellbeing compared with the running GPA. Goodman and Kruskal’s Gamma analysis revealed a moderately positive ordinal association between academic tolerance and multiclass wellbeing group (γ = 0.456). This suggests that higher academic tolerance is associated with a greater proportion of students experiencing higher overall wellbeing. Thus, our results are in agreement with Rathakrishnan et al. [13], who reported that academic self-perception was positively associated with life satisfaction among university students, suggesting that students’ perceptions of their ability to manage academic demands may be an important correlate of their overall wellbeing.

Students’ satisfaction with their relationships with others and their health emerged as the most influential factors out of the interpersonal and psychosocial factors explored in this study. Their relationships demonstrated the strongest positive ordinal association with overall wellbeing (γ = 0.505). Consistent with this finding, Ma et al. [14] have reported positive associations between interpersonal relationships, including relationships with parents, peers, and teachers, and psychological wellbeing among college students, further highlighting the importance of interpersonal experiences in students’ overall wellbeing. Among all 12 Likert-scale based variables examined, healthSatisfaction ranked third, demonstrating one of the strongest ordinal associations with wellbeing (γ = 0.460). Tavakoly Sany et al. [15] have reported that general health status was strongly associated with quality of life among university students, supporting the prominence of health-related perceptions in overall wellbeing. Together, these findings provide convergent evidence for the importance of interpersonal and health-related dimensions in student wellbeing.

Students’ satisfaction with the availability of money to meet their needs demonstrated a moderate ordinal association with overall wellbeing (γ = 0.377). The percentage distribution revealed that 31.8% of students reporting low money satisfaction (scores 1–4) belonged to the higher wellbeing group, compared with 64.4% of those reporting high money satisfaction (scores 7–9). These findings suggest that financial satisfaction is an important correlate of student wellbeing, although it may not be the sole factor determining overall happiness. In contrast, Richardson et al. [16] reported that greater financial difficulties among undergraduate students were associated with poorer mental health, including higher levels of depression and stress, and predicted poorer anxiety outcomes and global mental health over time. This apparent difference may reflect variations in the study populations, socioeconomic contexts, and sociocultural backgrounds. Furthermore, the relatively weaker association observed in the present study may suggest that other factors, such as social support and individual resilience, also contribute to the wellbeing of Sri Lankan undergraduates.

This study further applied interpretable machine learning techniques to classify psychological wellbeing status of undergraduates at the University of Peradeniya derived from self-reported overall happiness. XGBoost achieved the highest macro F1-score for binary classification, whereas random forest demonstrated the overall best performance for multiclass classification. These findings broadly resemble those of Rahman and colleagues [17], who found that random forest and adaptive boosting performed best among the algorithms evaluated for mental wellbeing classification across Southeast Asian universities. Similarly, Long and colleagues [12] identified random forest as the best-performing model for happiness and XGBoost for life satisfaction among Chinese university students. Although their relative rankings varied between the two classification approaches, SHAP analysis consistently identified academicTolerance, relationships, money_happiness, and healthSatisfaction as the four most influential features contributing to students’ overall wellbeing in both binary and multiclass classification.

### Strengths and Limitations

This study analyzed survey responses from 2,913 undergraduates at the University of Peradeniya. The data was collected using a newly developed, pilot-tested, and validated instrument covering multiple dimensions relevant to psychological wellbeing, including academic, social, financial, environmental, and personal factors. This multidimensional assessment enabled a broader characterization of student wellbeing than would be possible using psychological measures alone. Second, the application of multiple machine learning algorithms enabled assessment of the consistency of wellbeing classification across different modelling approaches while allowing potentially complex and non-linear relationships among predictors to be captured. Third, the integration of explainable artificial intelligence using SHAP enhanced model interpretability and facilitated identification of the factors contributing most strongly to model predictions. Importantly, the multiclass framework extended the analysis beyond binary classification by distinguishing lower, intermediate, and higher wellbeing, thereby providing a more nuanced representation of differences in student wellbeing.

Several limitations should nevertheless be acknowledged. First, the cross-sectional design precludes causal inference, and therefore, the identified predictors should be interpreted as predictive associations rather than causal determinants of psychological wellbeing. Second, the study relied predominantly on self-reported information, which may be susceptible to recall bias, response bias, and social desirability effects, although the use of a structured, pilot-tested, and validated instrument may have reduced some measurement-related concerns. Third, psychological wellbeing is dynamic and may vary with academic workload, examinations, financial circumstances, interpersonal experiences, and other changing life events. Consequently, a single assessment cannot fully capture temporal fluctuations in wellbeing or establish whether changes in the identified factors precede changes in wellbeing. Fourth, although rigorous internal validation and comparative model evaluation were undertaken, the absence of external validation in independent student populations limits conclusions regarding the transportability of the models beyond the Sri Lankan undergraduate student population. Differences in institutional context, socioeconomic conditions, academic systems, and student characteristics may affect model performance in other settings. Finally, machine learning models identify predictive patterns rather than underlying mechanisms. Accordingly, the predictors identified in this study should not be interpreted as definitive causal determinants but rather as potentially informative and actionable factors warranting further longitudinal and intervention-based investigation.

### Implications and Future Research

The findings suggest that efforts to improve psychological wellbeing among undergraduates should extend beyond individual psychological or clinical interventions and adopt a multidimensional approach. The importance of academic coping, goal achievement, health satisfaction, financial-related factors, interpersonal relationships, and residential and environmental conditions indicates that student wellbeing may be influenced by interconnected aspects of university life. Accordingly, universities could consider integrated wellbeing strategies combining academic support, goal setting and achievement programmes, peer and social support initiatives, health promotion activities, financial guidance, and improvements to student residential environments. Where appropriate, such initiatives could be complemented by counselling and other professionally supported mental-health services.

The findings may also have implications for early identification and targeted prevention. Machine learning models that can distinguish between lower, intermediate, and higher wellbeing may eventually support identification of students who could benefit from additional support, particularly when incorporated into ethically governed institutional wellbeing frameworks. However, such implementation should be approached cautiously, with appropriate attention to privacy, transparency, fairness, informed consent, and the risk of stigmatization. Predictive outputs should complement, rather than replace, professional assessment and student-centred decision-making.

Future research should build on these findings through longitudinal designs that can determine whether changes in the identified factors are associated with subsequent changes in wellbeing and can provide stronger evidence regarding temporal relationships. External validation across universities with different academic, demographic, socioeconomic, and cultural contexts is also essential before the models can be considered broadly generalizable. Future studies could further investigate whether model performance remains stable across relevant student subgroups and whether recalibration or context-specific model adaptation is required. In addition, qualitative research could provide deeper insight into the experiences and mechanisms underlying the associations identified by the machine learning models, particularly for factors such as academic tolerance, goal achievement, relationships, and residential environment.

## 5. Conclusions

This study demonstrates the potential of machine learning approaches to characterize and classify psychological wellbeing among university undergraduates using academic, social, financial, environmental, and personal variables. Extending the analysis from binary to multiclass classification provided a more differentiated assessment of lower, intermediate, and higher wellbeing levels, while comparisons across multiple algorithms demonstrated that predictive performance varied by modelling approach. The integration of SHAP enhanced interpretability by identifying the factors that contributed most strongly to individual model predictions.

Overall, the findings indicate that student psychological wellbeing is reflected in interconnected aspects of academic experiences, health, interpersonal relationships, financial circumstances, goal achievement, autonomy, and their living environments. Although the cross-sectional design precludes interpreting these factors as causal determinants, their contribution to model predictions highlights potential relevance for student wellbeing assessment and further research on intervention planning. Machine learning models may provide a complementary approach to identifying patterns relevant to student wellbeing and informing data-driven wellbeing university initiatives alongside established psychological assessment and support services.

These findings should be interpreted within the context of the Sri Lanka undergraduate student population and should not yet be assumed to generalize to other student populations. External validation across universities, regions, and culturally diverse populations is therefore essential, together with longitudinal studies examining whether the identified predictors are associated with subsequent changes in wellbeing. Future research should also examine whether interventions targeting the influential and potentially modifiable factors can produce measurable improvements in student wellbeing. Such work could support the development of validated, interpretable, and practically useful approaches to promoting psychological wellbeing in the higher education sector.

## Data Availability

Data is available from the corresponding author at a reasonable request.

## Acknowledgements

We gratefully acknowledge the financial support provided by the University Research Council, University of Peradeniya, Sri Lanka, through the Multidisciplinary Research Grant (Grant No. 370), which supported the successful completion of this research.

## Supporting Information

[S1 Text](https://docs.google.com/document/d/1oB3DtT84XhLvibKNwuXBlytn0ZQXuXi-P33ZEdqL9ag/edit?tab=t.0). Supplementary files for Explainable Machine Learning for Classification of PsychologicalWellbeing Status of Undergraduates of the University of Peradeniya, Sri Lanka.

## Notes

### Competing Interest Statement

The authors have declared no competing interest.

### Author Declarations

The Institutional Ethics Review Committe, Faculty of Arts, University of Peradeniya, Sri Lanka

